# Real-time fMRI-informed self-regulation of the ventromedial prefrontal cortex using positive episodic future thinking enhances motivation

**DOI:** 10.1101/2025.09.03.25334509

**Authors:** Yuan Zhang, Wenyi Dong, Xiangfeng Yan, Menghan Zhou, Keith M Kendrick, Dezhong Yao, Shuxia Yao, Benjamin Becker

## Abstract

Motivational deficits including anhedonia and avolition represent persistent challenges across psychiatric disorders, often resistant to conventional interventions. Here, we developed a novel brain-based precision intervention combining real-time fMRI neurofeedback with positive episodic future thinking to target the ventromedial prefrontal cortex (vmPFC), a region that critically regulates motivation via anticipated pleasure and value computation. A pre-registered randomized sham-controlled double-blind real-time fMRI trial (n=61) with personalized vmPFC targets demonstrated that the neurofeedback group rapidly acquired regulatory control over the vmPFC, leading to significantly higher activity of this region and enhanced anticipated pleasure and regulatory efficacy. Effects were maintained in the absence of feedback, underscoring the therapeutic potential. Network analyses revealed strengthened vmPFC-putamen connectivity following the training, reflecting network-level plasticity in motivational learning circuits. Results demonstrate rapid acquisition of vmPFC self-regulation and enhanced motivation through an ecologically-valid and pathway-specific strategy and underscore the therapeutic potential of personalized neurofeedback for motivational dysfunctions. (Clinical trials: NCT06787248)

## 1. Introduction

Motivation, particularly reward anticipation, is the fundamental process that shapes goal-directed behavior and supports adaptive functioning (Botvinick & Braver, 2015; Schacter, 2012; Schultz, 2015). Disruptions in motivational processes, particularly manifesting as symptoms of anhedonia, avolition, and apathy, are transdiagnostic symptoms across major psychiatric conditions, including major depressive disorder (MDD; Yang et al., 2022), psychotic disorders (Husain & Roiser, 2018; Strauss et al., 2021), and substance use disorders (Koob & Volkow, 2016). Among these, avolition and apathy often remain refractory to the established pharmacological treatments and non-invasive brain stimulation interventions that primarily target cortical surface regions (Bodén et al., 2021). These symptoms persist beyond acute episodes and significantly impair long-term outcomes, including social and occupational functioning (Craske et al., 2016; Strauss et al., 2021). Residual motivational deficits limit engagement with psychosocial interventions, impede recovery, and contribute to substantial healthcare costs (Vaccaro et al., 2025). The development of novel interventions that target the neurobiological basis of motivational deficits is thus a pressing priority (Bodén et al., 2021; Govil & Kantrowitz, 2025; Strauss et al., 2021).

Emerging evidence links avolition to transdiagnostic deficits in anticipated pleasure and episodic future thinking (EFT) - the ability to simulate personally relevant future events. Individuals with MDD and psychosis consistently show impairments in generating emotionally salient representations of rewarding future events, contributing to reduced anticipated pleasure and motivational salience (Frost & Strauss, 2016; Hallford et al., 2020; Yang et al., 2018). These deficits in EFT, particularly in the context of anticipated pleasure are associated with motivational impairments such as apathy and avolition, but independent of general cognitive dysfunction (Frost & Strauss, 2016; Hallford et al., 2020; Raffard et al., 2013).

Neuroimaging studies have begun to delineate a core network supporting anticipated pleasure and EFT, encompassing the medial prefrontal and medial temporal systems implicated in valuation, memory-based simulation, and goal representation (Andrews-Hanna, 2012; Schacter et al., 2007; Stawarczyk & D’Argembeau, 2015; Teghil et al., 2025). The ventromedial prefrontal cortex (vmPFC) has emerged as a central node within these domains and has been found consistently engaged during EFT (Benoit & Schacter, 2015; Ciaramelli et al., 2021a) while damage to this region critically impairs future-oriented, self-relevant simulation (Ciaramelli et al., 2021a) and future reward valuation (Ciaramelli et al., 2021b). Accumulating evidence additionally indicates that the vmPFC is associated with value-based decision-making (Zangemeister et al., 2019) and reward valuation (Doré et al., 2017) and supports EFT via its contribution to positive foresight (Benoit et al., 2014; Hu et al., 2024). Together these results indicate that this region plays a key role in internal prospection, motivation, goal-directed behavior, and affective prediction (Zhou & Becker, 2025).

Real-time functional magnetic resonance imaging (rt-fMRI) neurofeedback (NF) training enables individuals to gain volitional control over specific brain regions by providing real-time feedback on neural activity. Prior studies have demonstrated that rt-fMRI NF training can selectively modulate activity in target regions (Dudek & Dodell-Feder, 2021; Linhartová et al., 2019) or the communication between regions (Morgenroth et al., 2020; Zhao et al., 2019), with region-specific changes in affective, reward, and motivational outcomes (Li et al., 2018; Martz et al., 2020; Yao et al., 2016). Initial work has explored the vmPFC as target region in healthy participants (Garrison et al., 2021; Mayeli et al., 2020), and a recent pilot study examined vmPFC-based NF in MDD (Aupperle et al., 2024). The MDD study reported increased vmPFC activation across training runs and a trend-level improvement in symptoms when participants used positive EFT as a regulation strategy. Despite the promising results, the small sample (n=6), lack of a control group, and absence of behaviorally relevant endpoints limit the study’s conclusions.

Against this background, the present study aimed to determine the feasibility of a positive EFT strategy in regulating the vmPFC activation using a pre-registered randomized double-blind, sham feedback-controlled design in a well-powered sample of healthy participants. Based on prior literature and the objectives of our study, we pre-registered the trial to assess the efficacy of real-time fMRI neurofeedback training at both the neural and behavioral levels (https://clinicaltrials.gov/study/NCT06787248). Our primary neural outcome was a training-induced increase in the left vmPFC in the NF training (NFT) group compared to the sham control (CTR) group. Participants in the CTR group received feedback from a region not implicated in positive anticipation (the left primary motor cortex). Secondary outcomes focused on between-group differences in training-induced enhancement of anticipated pleasure, assessed via self-report ratings, and motivational effort, assessed via the Effort Expenditure for Rewards Task (EEfRT). Additional exploratory outcomes included: (1) the maintenance of increased vmPFC activation and anticipated pleasure during a transfer run without real-time NF, and (2) changes in vmPFC functional connectivity (FC) with brain regions involved in regulatory control and reinforcement learning, as assessed through fMRI during and after the training.

## 2. Materials and Methods

### 2.1. Participants

61 healthy participants (31 males, mean age=22.61, SD=2.15) were enrolled in a pre-registered randomized double-blind, sham feedback-controlled parallel-group trial with n=31 (16 males, 21.45, SD=2.06) randomly assigned to receive continuous rt-fMRI NF from the left vmPFC in the NFT group and n=30 (15 males, 21.77, SD=2.27) assigned to the CTR group receiving sham feedback from a region not typically engaged in positive future thinking or motivational functions (left primary motor cortex, M1) (similar approach see Zhao et al., 2019 and additional analyses provided in supplements and Table S3). Data from 8 participants were excluded due to excessive head movement (rotation or motion>3mm; 5 participants), study withdrawal (1 participant) or MRI data quality (2 participants) leaving n=53 participants for final analyses (NFT group: 27 participants, 12 males, mean age=21.41 years, SD=2.19; CTR group: 26 participants, 14 males, mean age=21.65 years, SD=2.11; details see CONSORT in Fig. S1). All participants gave written informed consent and did not have a current or history of mental or neurological disorder and psychotropic substance use. All experimental procedures were approved by the local ethical committee at the University of Electronic Science and Technology of China and adhered to the latest version of the Declaration of Helsinki. This study was preregistered on clinicaltrials.gov (<u>NCT06787248</u>) with NF training-induced changes in vmPFC activity as the primary outcome and changes in anticipated pleasure (ratings) and motivational effort in EEfRT as the secondary outcomes.

### 2.2. Experimental Procedure

#### 2.2.1. Assessment of confounders

Before the experiment participants completed a series of validated Chinese versions of psychometric questionnaires assessing affective state and factors related to imagination (Figure 1A), including Beck Depression Inventory-II (Beck et al., 1996), State-Trait Anxiety Inventory (STAI; Spielberger et al., 1971), Vividness of Visual Imagery Questionnaire (VVIQ; Marks, 1973), Behavioral Inhibition System and Behavioral Activation System Scale (BIS/BAS; Carver and White, 1994), and Zimbardo Time Perspective Inventory (ZTPI; Zimbardo & Boyd, 1999). Unspecific affective changes during NF training were assessed using the Positive and Negative Affect Schedule (PANAS; Watson et al., 1988) and State Anxiety Inventory administered before and after the training task.

**Figure 1.**
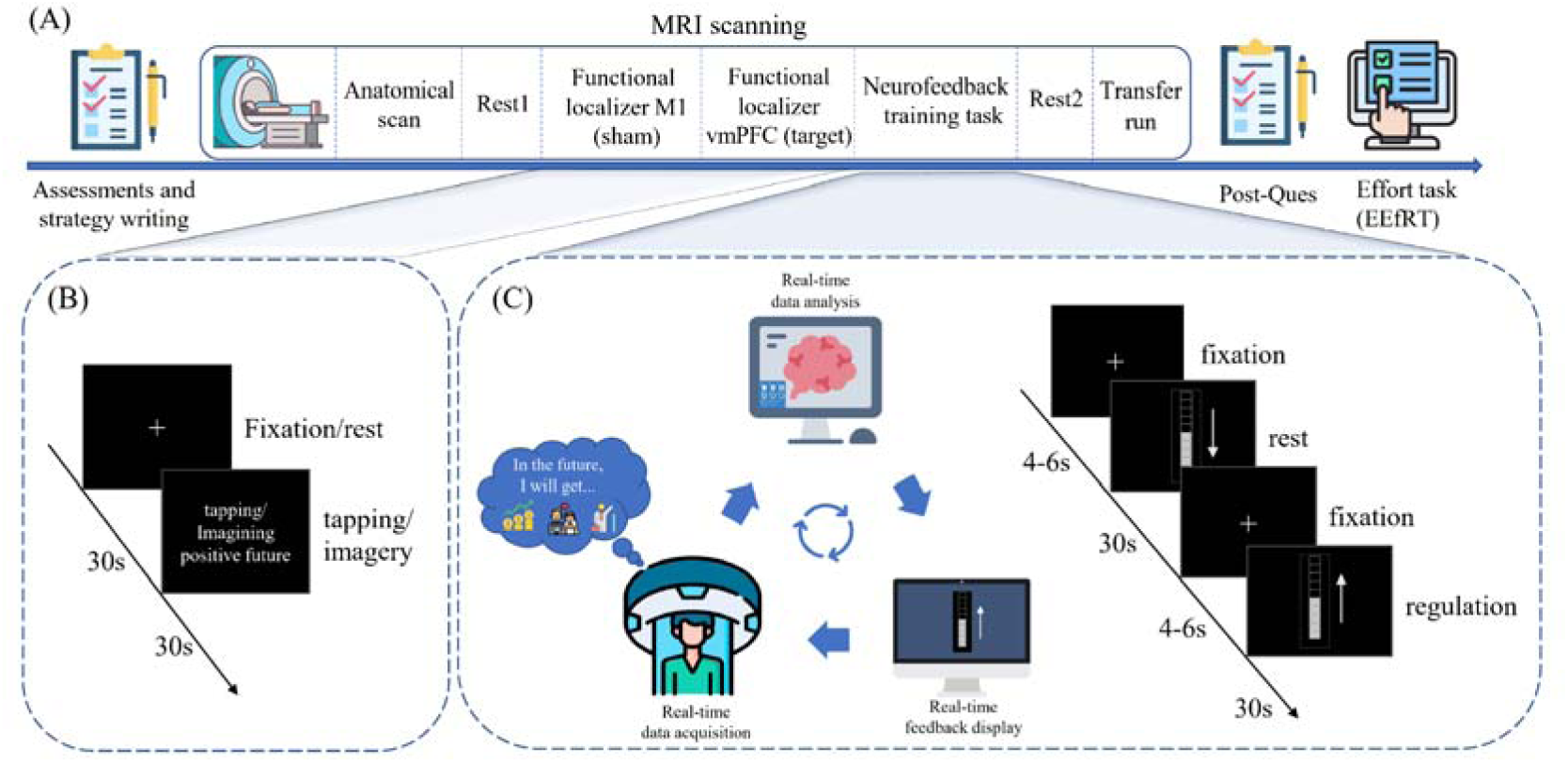
(A) Experimental protocol. Experimental paradigms for the funtional localizer tasks (B) and neurofeedback training task (C).

#### 2.2.2. NF training protocols and regulation strategies (primary outcome)

Based on previous studies demonstrating that positive future imagination enhances motivation and robustly engages the vmPFC (Ji et al., 2021; Schacter et al., 2017; Thakral et al., 2017), we utilized this mental process to develop individualized neuromarkers and regulation strategies. In a pre-fMRI training participants were requested to vividly imagine three most joyful events that could occur in their future, drawing upon their own experience or using events from a reference list (ref. Speer et al., 2021). The subsequent rt-fMRI NF protocol (see Figure 1A) consisted of (1) two localizer runs to determine individualized localization of the vmPFC involved in future positive thinking as well as the M1 during which participants were required to imagine the joyful future events or tap their fingers, respectively; (2) random assignment to the training or sham control condition for four runs NF training during which participants aimed at enhancing the activity of the target region with NF provided from the vmPFC (NFT) or M1 (CTR); and (3) one transfer run without rt-fMRI NF to test the maintenance of the training success in the absence of feedback. The training was embedded in two resting-state fMRI runs before (Rest1) and immediately after the NF training (Rest2) to explore training-related FC changes on the network level using an eyes-open acquisition protocol.

For all participants the experiment started with two localizer paradigms. The M1 localizer paradigm instructed participants to tap their right index finger approximately once per second (Figure 1B; details see Table S3). The vmPFC localizer task required all subjects to imagine the three most joyful events they wrote in the positive imagination blocks and to keep relaxed in the rest blocks. The localizer run was also used as a pre-training baseline.

The subsequent training runs consisted of alternating five regulation and five rest blocks, each with a 30s duration. In line with previous studies (Yao et al., 2016; Zhang et al., 2023; see Figure 1C), participants were informed that the direction of an arrow displayed along the continuous feedback bar indicated the specific condition (i.e. an up arrow indicates regulation and a down arrow indicates rest blocks). For performing the regulation strategy, all participants were instructed to up-regulate brain activity via “positive EFT” during regulation blocks and to relax during rest blocks. The last transfer run was identical to the prior training run, yet without feedback being provided, and served to determine the maintenance of regulatory control in the absence of NF (Yao et al., 2016; Young et al., 2017; Zhang et al., 2023). Before the training, participants were informed that the NF would be delayed due to the nature of the hemodynamic response and that gaining control may take some time.

#### 2.2.3. Motivational effects of training: anticipated pleasure and motivational effort (secondary outcomes)

Effects of training on motivation were evaluated at two levels: subjective experience of anticipated pleasure during training, and motivational effort assessed after the training. At the level of subjective experience, two dimensions were assessed following each of the four training runs and the transfer run: (1) anticipated pleasure (“How pleasant do you feel about the upcoming events?”; 1=not at all, 9=very pleasant), and (2) regulatory efficacy (“How effective was the regulation strategy you used?”; 1=not at all, 9=very effective) using self-report rating scales. The motivational effort was assessed behaviorally using the EEfRT, administered after the NF training. In this modified version of the EEfRT (Gill et al., 2023; Ha et al., 2022; Treadway et al., 2009), participants chose between an ’easy task’ and a ’hard task’, each trial associated with one of three reward probabilities: high (88%), medium (50%), or low (12%). The ‘easy task’ always yielded a fixed reward of 1 yuan, while the ‘hard task’ offered a larger reward ranging from 1.2 to 4.2 yuan. During the decision phase, participants had 5s to make their choice. The task execution effort varied accordingly: the ‘easy task’ required 21 button presses within 7s, while the ‘hard task’ required 100 button presses within 30s. The feedback consisted of individual task performance and the reward amount was displayed within 2s. The total task duration was 10min, details see Figure S2.

### 2.3. Image Data Acquisition

All images were acquired on a 3T, GE Discovery MR750 scanner (General Electric Medical System, Milwaukee, WI, USA). High-resolution whole-brain volume T1*-weighted images were first collected with a 3D spoiled gradient echo pulse sequence (repetition time, 6 ms; echo time, minimum; flip angle, 12◦; field of view=256×256 mm; acquisition matrix, 256×256; thickness, 1 mm; number of excitations, 2; 172 slices). Functional images were acquired using a T2*weighted echo-planar imaging pulse sequence (repetition time, 2000 ms; echo time, 30 ms; slices, 32; thickness, 3.4 mm; gap, 0.6 mm; field of view, 220×220 mm; resolution, 64×64; flip angle, 90◦).

### 2.4. Data analyses

#### 2.4.1. Online real-time fMRI NF analyses

The rt-fMRI platform and protocols were identical to our previous studies (Yao et al., 2016; Zhang et al., 2023), real-time data processing was based on Turbo Brain Voyager (TBV, Version 4.0, Brain Innovation B.V., Maastricht, Netherlands). To facilitate personalized localization of the vmPFC we combined individualized anatomical and functional data. The anatomical images were transferred to TBV and preprocessed including brain extraction, inhomogeneity correction, and MNI normalization, and overlayed with a left medial PFC mask extracted from the Yeo-17 atlas (Yeo et al., 2011; see Figure S3). An 8-mm sphere was defined centering on the maximally activated voxel within the left vmPFC anatomical mask during positive future imagination relative to the rest condition in the localizer task. For the sham control group, an 8-mm sphere was defined centering on the maximum activity in the M1 during the finger tapping task. During NF training, data were preprocessed online using the TBV 4.0 including 3D motion correction and drift removal. The thermometer visualization indicative of the BOLD signal level of the target/control ROI was created by TBV 4.0 and then displayed via E-prime 3.0 in real-time.

#### 2.4.2. Offline imaging data analyses

SPM12 (Wellcome Department of Cognitive Neurology, London, UK, http://www.fil.ion.ucl.ac.uk/spm) was used for offline image processing. After removing the first 5 volumes, the remaining images were preprocessed including head-motion correction, co-registration of the mean functional image and the T1 image, normalization to the standard Montreal Neurological Institute (MNI) space, and smoothing with a Gaussian kernel (8 mm full-width at half maximum; FWHM). For the vmPFC localizer task, the first-level design matrix included 2 regressors (positive imagination and rest) and the 6 head motion parameters convolved with the canonical hemodynamic response function (HRF). For each training and transfer run, the first-level design matrix included 2 regressors (regulation and rest) and the 6 head motion parameters convolved with the canonical HRF. Contrast images between the aforementioned 2 conditions were created for the vmPFC localizer run (‘positive imagination>rest’), each training run, the transfer run, and across 4 training runs (‘regulation>rest’) for each participant.

For the second level analysis, we initially conducted a series of quality assurance and control analyses. For the vmPFC localizer task, a one-sample t-test on the contrast of ‘positive imagination>rest’ was first performed to validate vmPFC engagement during the functional localizer on the group level. A two-sample t-test on the same contrast in the vmPFC localizer task was then conducted to determine whether there was a group difference before the NF training. For the NF training task, a one-sample t-test was first applied across 4 training runs at the whole-brain level to examine general neural engagement during the NF training using the contrast of ‘regulation>rest’ for the NFT and CTR groups separately and exploratory two-sample tests were used to explore group differences. At the whole-brain level, results were corrected using the False Discovery Rate peak-level correction (*p*_FDR_<0.05) and only clusters larger than 10 voxels were reported.

#### 2.4.3. Analysis of primary outcomes (neural efficacy)

Effects of training on the neural level were assessed using ROI analysis of the target region in the NFT and CTR groups (see also Yao et al., 2016; Zhang et al., 2023). The target ROI was determined using the peak-coordinate within the left vmPFC in the localizer task (MNI: X=-3, Y=59, Z=-13; 8-mm sphere) as well as an independently defined atlas-based structural mask of the vmPFC (i.e. Frontal_Med_Orb_L in the AAL3; Rolls et al., 2020). Training-induced group differences were examined using extracted parameter estimates from the vmPFC on the contrast of ‘regulation>rest’ for each training run. A repeated-measures ANOVA was performed on vmPFC parameter estimates with run as a within-subject factor (run 1–4) and group (NFT vs. CTR) as a between-subject factor (using SPSS version 22, SPSS Inc., Chicago, IL, USA) during NF training. Two-sample t-tests were conducted to exclude group differences in vmPFC parameter estimates before NF training (localizer run) and to examine group differences in the maintenance of the regulation ability (transfer run) in the absence of feedback.

#### 2.4.4. Analysis of secondary outcomes (behavioral efficacy)

Repeated-measures ANOVAs were conducted to analyze behavioral NF training effects on rating scores of anticipated pleasure and the efficiency of the regulation strategy used during 4 training runs. Bonferroni correction was applied for multiple comparisons in post-hoc analyses disentangling significant main or interaction effects. A repeated-measures ANOVA was conducted on the percentage of ‘hard-task choice’ in the EEfRT following NF training. Brain-behavior correlations were tested using Pearson correlations given normal distribution of the data. Additionally, group differences in potential confounders were examined using two-sample t-tests. The mood changes assessed by PANAS and SAI were examined using mixed ANOVAs for NF and CTR groups, separately. In all cases, p<0.05 (two-tailed) was considered as significant.

#### 2.4.5. Exploratory analyses: Task-based and resting-state functional connectivity analyses

We conducted a psychophysiological interaction (PPI) analysis using the gPPI toolbox (McLaren et al., 2012) to examine task-based FC changes between the vmPFC and other brain regions during the NF training task (contrast: ‘regulation>rest’). The seed region was also defined as an 8-mm sphere centered on the peak-coordinate within the vmPFC in the localizer task. A one-sample t-test was performed for the NFT and CTR groups, respectively. The group difference in the task-based FC during NF training was analyzed by a two-sample t-test.

For the resting-state data standard processing procedures were employed including removal of the first 5 volumes, the remaining images were preprocessed with slice timing correction, realignment, and spatial normalization. The 6 head-motion parameters (using the Friston 24-parameter model), global signal, white matter, and cerebrospinal fluid as covariates were regressed to exclude potential confounding artifacts (Fox et al., 2005). The normalized functional images were resampled using a 3mm×3mm×3mm resolution. Next, these images were spatially smoothed (8mm FWHM), linearly detrended and filtered using a bandpass filter of 0.01–0.08. The resting-state functional connectivity (rsFC) analysis was conducted to examine rsFC changes between the vmPFC and other regions induced by NF training for each group and between these two groups. The seed region was also defined in the vmPFC localizer run (MNI coordinates: X=-3, Y=59, Z=-13; 8-mm sphere). The time series of each subject was extracted from the seed ROI separately for each resting scan. The FC map was generated based on a voxel-wise correlation analysis between the vmPFC and other voxels in the whole brain and then converted into a Z-map using Fisher’s z transformation. Furthermore, we conducted a flexible factorial ANOVA model to examine rsFC changes of time-specific effects for each group by subtracting the FC in Rest1 before NF training from the FC after training in Rest2 for NFT and CTR groups, separately.

## 3. Results

### 3.1. Demographics and questionnaires

The NFT and CTR groups displayed comparable demographics, affective states, and personality traits (all *ps*≥0.163; see Table S1). Mixed ANOVAs on scores of the PANAS-positive and SAI also revealed no significant main effect and interaction effect, arguing against nonspecific effects of training (all *ps*≥0.216; Table S2). There was no significant main effect of group and group×timepoint interaction effect in the scale of PANAS-negative (Table S2), but a significant main effect of timepoint was observed (F(1,51)=6.129, *p*=0.017, ηp^2^=0.107), indicating that negative mood in both groups improved following the NF training.

### 3.2. Primary and secondary outcomes

#### 3.2.1. Primary outcomes: neural efficacy and its maintenance

A two-sample t-test on the vmPFC activity in the localizer task showed that the groups did not differ in vmPFC engagement during positive future imagination before the NF training (t=1.605, *p*=0.115, Cohen’s d=0.442). A mixed ANOVA with factors of run and group on the vmPFC activity during training revealed a significant main effect of group (F(1,51)=5.309, *p*=0.025, ηp^2^=0.094), with stronger activation of the vmPFC in the NET compared to the CTR group (Figure 2A). No significant main effect of run (F(3,153)=0.690, *p*=0.560, ηp^2^=0.013) or group×run interaction effect (F(3,153)=0.250, *p*=0.861, ηp^2^=0.005) was observed.

**Figure 2.**
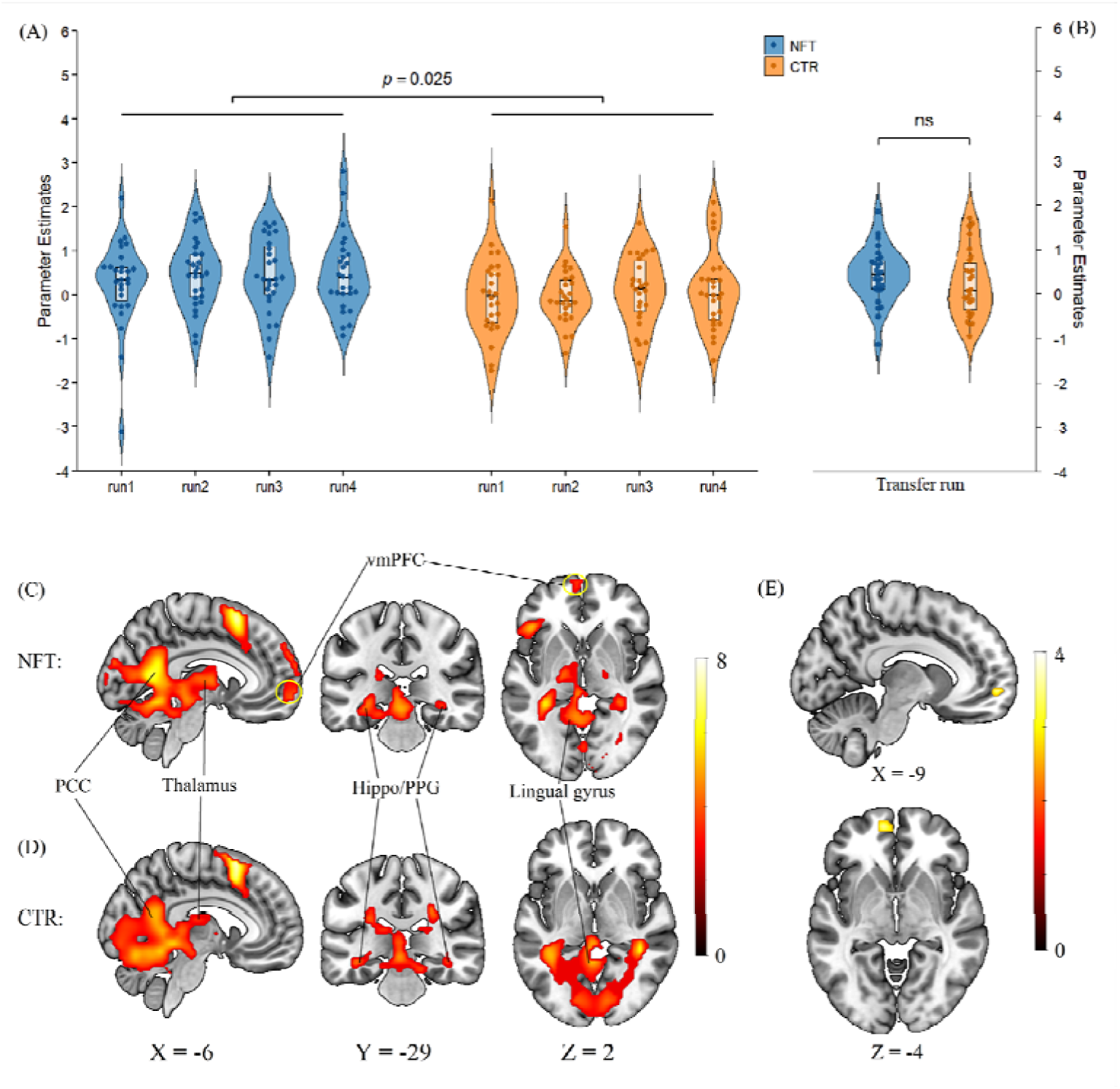
(A) Left ventromedial prefrontal cortex (vmPFC) activity in the regulation blocks relative to rest blocks over the 4 training runs and (B) in the transfer run. Error bars represent standard error. (C) Left vmPFC activity in the whole-brain level analysis across the 4 training runs in the neurofeedback training (NFT) and (D) the sham control (CTR) groups (*p*_FDR_<0.05). (E) Left vmPFC activity in the whole-brain level analysis in the NFT compared with the CTR group using a small volume correction. PCC: Posterior cingulate cortex; Hippo: Hippocampus; PPG: Parahippocampal gyrus.

Notably, the robustness of the results was further validated using an independent and regional-specific structural mask of the vmPFC in the AAL3 with a similar mixed-ANOVA replicating a significant main effect of group (F(1,51)=7.628, *p*=0.008, ηp^2^=0.130), but no significant main effect of run (F(3,153)=0.205, *p*=0.893, ηp^2^=0.004) and group×run interaction effect (F(3,153)=0.161, *p*=0.922, ηp^2^=0.003). While the NFT group continued to show higher vmPFC engagement during the transfer run, group differences failed to reach significance (ROI based on the localizer, t=0.947, *p*=0.348, Cohen’s d=0.260, Figure 2B; structural mask, t=1.422, *p*=0.161, Cohen’s d=0.398).

#### 3.2.2. Confirmatory whole-brain analyses

A one-sample t-test on the localizer data for both groups confirmed the vmPFC engagement during positive future thinking (MNI coordinates: X=-3, Y=59, Z=-13) and the engagement of other regions involved in this process (e.g., posterior cingulate cortex (PCC), hippocampus, parahippocampal gyrus (PPG), p_FDR_<0.05; see Table S3), while the results confirmed the lack of M1 engagement and a two-sample t-test confirmed the absence of pre-training group differences before NF training (p_FDR_<0.05).

Exploring the training-related activity on the whole brain level using a one-sample t-test on contrast ‘regulation>rest’ across 4 training runs showed significant activity in the vmPFC, PCC, PPG, putamen, and other frontal-temporal regions in the NFT group (*p*_FDR_<0.05; Figure 2C and Table S4). In contrast, the CTR group did not display engagement of the vmPFC but the PCC, PPG, and other frontal-temporal regions (*p*_FDR_<0.05; Figure 2D). A whole brain voxel-level analysis using paired t-tests on the contrast ‘regulation>rest’ reveal no significant group differences across 4 NF training runs or in the transfer run (*p*_FDR_<0.05), but a confirmatory analysis focused on the vmPFC using small volume correction revealed significant group differences on the regional level (*p*_FDR_=0.019, t=3.74, k=19; Figure 2E). For the transfer run, no significant group differences were observed.

#### 3.2.3. Secondary outcome: motivational experience and maintenance

Examining the secondary outcome of anticipated pleasure using a mixed ANOVA with run as within-subject factor and group as between-subject factor revealed a significant main effect of group (F(1,51)=11.577, *p*=0.001, ηp^2^=0.185; Figure 3A), indicating a higher anticipated pleasure in the NFT group. No significant main effect of run (F(3,153)=2.097, *p*=0.103, ηp^2^=0.039) and group×run interaction effect (F(3,153)=0.566, *p*=0.638, ηp^2^=0.011) was observed. For regulatory efficiency ratings a corresponding ANOVA revealed a significant main effect of group (F(1,51)=16.476, *p*<0.001, ηp^2^=0.244; Figure 3B), reflecting a higher experienced control in the NFT group. No significant main effect of run (F(3,153)=1.228, *p*=0.254, ηp^2^=0.026) and group×run interaction effect (F(3,153)=1.729, *p*=0.163, ηp^2^=0.033) were observed. Higher anticipated pleasure and regulatory efficiency in the NFT group were maintained during the transfer run as revealed by two-sample t-tests (anticipated pleasure; t=2.915, *p*=0.005, Cohen’s d=0.797; efficiency, t=3.019, *p*=0.004, Cohen’s d=0.828; Figure 3C). No differences were observed during the vmPFC localizer run for anticipated pleasure (t=1.039, *p*=0.304, Cohen’s d=0.286) and regulatory efficiency (t=0.942, *p*=0.351, Cohen’s d=0.260), confirming training-related effects.

**Figure 3.**
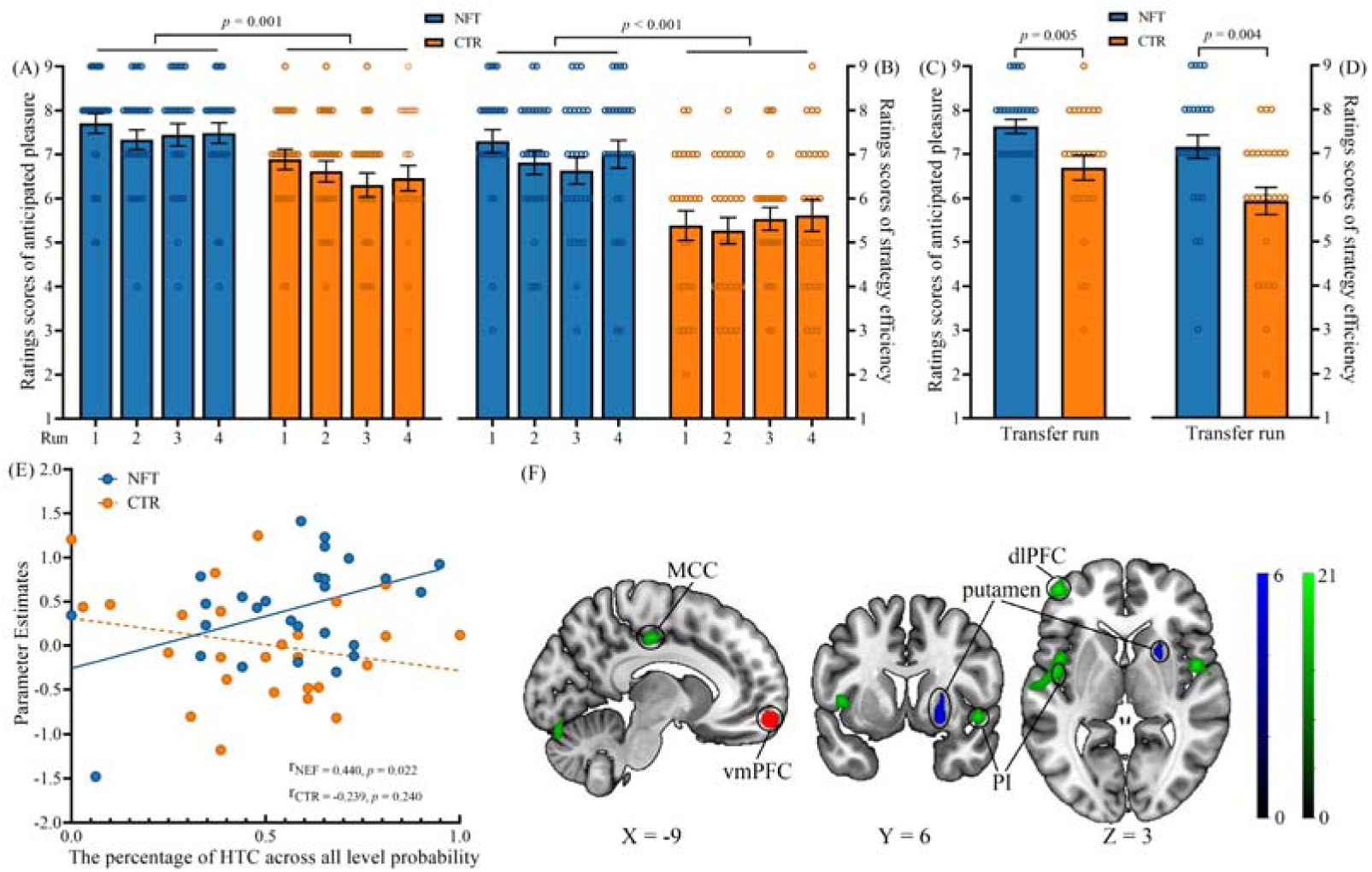
Rating scores of anticipated pleasure and the regulatory efficiency for neurofeedback training (NFT) and sham control (CTR) groups over the 4 training runs (A-B) and during the transfer run (C-D). Error bars represent standard error. (E) Pearson correlations between the percentage of ‘hard-task choices’ (HTC) in the Effort Expenditure for Rewards Task and the left ventromedial prefrontal cortex activity across 4 training runs in the NFT and CTR groups. (F) Regions showed increased task-based functional connectivity (the blue; *p*_FDR_<0.05) during NF training and increased resting-state functional connectivity (the green; *p*_uncorrected_<0.001) after NF training using the ventromedial prefrontal cortex (vmPFC; the red) as the seed region. dlPFC: dorsolateral prefrontal cortex; MCC: middle cingulate cortex; PI: posterior insula.

#### 3.2.4. Secondary outcome: motivational effort

In the EEfRT, a repeated-measures ANOVA with reward probability as within-subject factor and group as between-subject factor on the percentage to ‘hard-task choice’ revealed a significant main effect of reward probability (F(2,102)=29.164, *p*<0.001, ηp^2^=0.364), indicating that participants had a stronger trend to select ‘hard-task’ with requirement of making more effort when probability to obtain a monetary reward got higher, but no significant main effect of group (F(1,51)=1.009, *p*=0.320, ηp^2^=0.019) or interaction effect of group×probability (F(2,102)=0.221, *p*=0.802, ηp^2^=0.004).

### 3.3. Exploratory analyses

#### 3.3.1. Exploratory results: neural training effects and motivational effort

We conducted Pearson correlation analyses to further determine potential associations between vmPFC activity related to NF training and the motivational tendency to pay a higher effort to get a higher reward across all probability levels. Results revealed a significant positive correlation in the NFT (Pearson r=0.440, *p*=0.022; Figure 3D) but not in the CTR group (Pearson r=-0.239, *p*=0.240; Figure 3D). The correlation difference between groups was also examined using the Fisher z-transformation test and revealed a significant difference between the NFT and CTR groups (Fisher z-scores=2.454, *p*=0.014).

#### 3.3.2. Exploratory results: NF-induced network-level changes

We next explored whether the training altered the coupling of the vmPFC with other brain regions using a PPI analysis based on contrast ‘regulation>rest’ in the NFT group (ref. Caria et al., 2007; Yao et al., 2016). The NFT group displayed enhanced vmPFC-right putamen connectivity during NF training at the whole brain level (MNI coordinates: X=24, Y=11, Z=-7; t=6.05, k=60; Figure 3F; *p*_FDR_<0.05), while no connectivity differences were observed in the CTR group. However, no significant differences in task-based FC changes were found between NFT and CTR groups (*p*_FDR_<0.05).

A flexible factorial ANOVA model was conducted to examine the rsFC changes of each group and group differences induced by NF training. There was no surviving cluster after FDR correction in the NFT and CTR groups on contrast of ‘Rest2>Rest1’ or the corresponding group comparison (i.e. NFT_Rest2>Rest1_>CTR_Rest2>Rest1_; *p*_FDR_<0.05). However, an exploratory analysis in the NFT group showed stronger rsFC between the vmPFC and a series neural networks, including the left dorsolateral PFC (dlPFC), left middle cingulate cortex (MCC), the bilateral posterior insula (PI), the left angular gyrus (AG) and the left lingual gyrus (*p*_uncorrected_<0.001; Figure 3F and Table S5).

## 4. Discussion

The present pre-registered double-blind sham-controlled trial aimed to determine whether positive EFT can serve as strategy to gain volitional self-regulation of the vmPFC and, in turn, enhance motivational processes. During NF training, the NFT - compared with the CTR - group demonstrated significantly higher vmPFC activity and increased motivation in terms of enhanced experience of anticipatory pleasure and regulatory efficacy. A trend level of stronger vmPFC activity for the NFT group compared to the CTR group was also found in the transfer run indicating that regulatory control is maintained to some extent in the absence of feedback. Importantly, anticipated pleasure and regulatory efficacy remained statistically significant higher during the transfer run in the NFT – compared to the CTR – group, underscoring maintained motivational effects. On the network level the NFT group showed increased functional communication between the vmPFC and specifically the putamen, a region critically involved in both motivational processes and regulatory control. Exploratory analyses showed training-induced changes in the functional architecture of the brain such that the NF group exhibited enhanced intrinsic communication of the vmPFC and with core regions of the motivational and cognitive control networks. While no group differences were observed in motivational effort, a positive association between vmPFC activity and the tendency for a ‘hard-task choice’ was observed specifically in the NFT group.

With respect to the neural training efficacy (primary outcome) results from region and voxel-based analyses convergently demonstrated that participants in the NFT group exhibited a stronger activation of the vmPFC target during the training compared to the CTR group. Higher activity was observed across all four training runs, reflecting the feasibility of positive EFT as the regulation strategy in allowing rapid and consistent regulatory control over vmPFC activation. The rapid acquisition of neural control is in line with recent studies employing strategies that functionally align with the target region and allow participants to gain regional-specific neural self-regulation, including value-based thinking to control the vmPFC, interoception to control the anterior insula and reappraisal to control the lateral prefrontal activation (Mayeli et al., 2020; Tsang et al., 2025; Zhang et al., 2023). Notably, both groups in the present study received the same strategy instructions, underscoring the critical contribution of the NF in gaining rapid and consistent regulatory control. Important to the translation of the findings into real-world and clinical applications, the NFT group maintained – although non-significantly - higher activation of the vmPFC during the transfer run in the absence of feedback. A more robust maintenance effect may be achieved by optimizing the NF training protocol such as including more NF training runs in one visit or splitting into different visits (Arns et al., 2014; Thibault et al., 2017, 2018). Together with a recent pilot study demonstrating the feasibility of positive EFT-based NF targeting the vmPFC in six individuals with suicidal ideation and depression (Aupperle et al., 2024), the findings from our pre-registered, double-blind, sham-controlled study with a comparatively large sample (N=61) provide convergent evidence supporting the neurobiological and clinical potential of positive EFT-based vmPFC self-regulation for mood and affective disorders.

Examination of the secondary behavioral outcomes confirmed that regulatory control in the NFT group led to greater anticipated pleasure and higher perceived regulatory efficacy compared to the CTR group. Consistent with previous findings (Heise et al., 2025; Renner et al., 2019), positive EFT elicited pleasant experiences and anticipated pleasure in both groups. However, the combination of EFT with rt-fMRI NF further amplified anticipated pleasure specifically in the NFT group. Importantly, the training effects on anticipated pleasure and perceived control in the NFT group could be maintained during the transfer run in the absence of feedback compared to the CTR group, with medium to large effect sizes for these subjective experience measures. In contrast, we did not find strong evidence for effects on motivational effort, although the NFT group tended to exert greater efforts to get a higher reward across all probability levels.

Exploratory correlation analyses further revealed an association between higher vmPFC activation and higher motivational effort specifically in the NFT group significantly different from the correlation pattern in the CTR group. Anticipated pleasure and perceived control are central components of motivation, with the former driving reward expectancy and the latter supporting goal-directed setting through a sense of agency. Deficits in these domains are well-documented in mental disorders and are key contributors to motivational impairments (Frost & Strauss, 2016; Hallford et al., 2020; Yang et al., 2018). The vmPFC plays a crucial role in integrating these signals - encoding subjective value, perceived agency, and affective prospection (Benoit et al., 2014; Hu et al., 2024; Zhou & Becker, 2025) - to guide decision-making and goal-directed behavior. In this context, our findings not only demonstrate a causal role of the vmPFC in supporting EFT and its association with motivational processes, but also highlight the potential of vmPFC-targeted NF training to modulate these mechanisms and inform translational interventions for clinical populations with motivational deficits (in support of Aupperle et al., 2024).

An exploratory analysis of training-induced changes of vmPFC communication with other brain systems showed enhanced coupling between the target region and the putamen and preliminary evidence for changes in the intrinsic communication of the vmPFC with lateral frontal, mid-cingulate and posterior insula – yet not core DMN regions - in the NFT group. The putamen and caudate, key components of the dorsal striatum, play central roles in learning and reward processing (Kreitzer & Malenka, 2008; Valjent & Gangarossa, 2021), particularly in reinforcement learning and reward anticipation (Daniel & Pollmann, 2014; Ghandili & Munakomi, 2025). Prior empirical studies (Young et al., 2018; Zhao et al., 2021) and theoretical frameworks (Lubianiker et al., 2022; Sitaram et al., 2017) have indicated a critical role of the putamen in NF training, with increasing evidence supporting this association across feedback modalities (e.g., EEG, fMRI), study designs, and populations (Emmert et al., 2016; Enriquez-Geppert et al., 2013; Ninaus et al., 2015; Zhao et al., 2021). In clinical contexts, dysfunction of the putamen has been linked to impaired reward anticipation and motivational deficits such as anhedonia and avolition (Bore et al., 2024; Daniels et al., 2025; Zimmermann et al., 2019). Within this framework, the observed increase in functional connectivity between the vmPFC and the putamen in the NFT group may reflect a dual mechanism of enhanced learning of regulatory control and facilitated positive reward anticipation.

A highly exploratory analysis revealed that within the NFT group the training induced changes in vmPFC communication with a left lateralized network encompassing the dlPFC, MCC, the bilateral left lingual gyrus and bilateral posterior insula (*p*_uncorrected_<0.001; Table S5), yet – in contrast with a previous NFT study targeting the vmPFC – not the DMN (Mayeli et al., 2019). Findings may reflect that the training-induced changes on the network level may vary depending on the specific vmPFC subregion and mental process targeted and that the vmPFC as a core region of the DMN can be retrained on the network level. The network found in the present study shows engagement in a range of general mental processes ranging from cognitive control and self-regulation (dlPFC; Hare et al., 2009; Rudorf & Hare, 2014; Schmidt et al., 2018), to performance monitoring (MCC, Di Cesare et al., 2021; Shenhav et al., 2013) and interoceptive signaling (Critchley et al., 2004; J. Li et al., 2019), which together may further confirmed the intrinsic reconfigurations of brain networks induced by NF training, although these effects may vary given different target regions and regulation strategies(Marins et al., 2019; Misaki et al., 2018; Yao et al., 2016).

Our findings require interpretation in the context of limitations. First, effects on the secondary outcome of motivational effort as assessed by the EEfRT failed to reach significance, and results may reflect dissociable neural representations of anticipatory pleasure and effort, a question that may be evaluated by future studies. Second, anticipated pleasure and regulatory efficiency were assessed at the end of each run rather than immediately following each regulation block and continuous online assessment may provide more detailed results. Lastly, results of the exploratory correlation and network analyses require cautious interpretation and replication in future studies.

In summary, the present pre-registered randomized sham-controlled trials provide convergent evidence that a brief rt-fMRI NF session targeting the vmPFC, and combined with positive EFT, enables healthy individuals to volitionally increase vmPFC activity and maintain this modulation beyond the training context. The neural regulation success enhanced anticipated pleasure and experienced regulatory control, establishing a direct link between vmPFC-targeted neurofeedback, and core processes underlying motivation. These findings underline the translational potential of vmPFC-based NF interventions as an innovative approach for targeting motivational deficits, with particular promise in conditions characterized by anhedonia and avolition.

## Data Availability

All data produced in the present study are available upon reasonable request to the authors.

## Acknowledgements

This work is supported by the National Natural Science Foundation of China (NSFC 82271583), the Ministry of Science and Technology of China (STI 2030–Major Projects 2022ZD0208500), the Hong Kong University Grants Council (GRF 17615525), the University of Hong Kong seed funding and start-up schemes (2407102536).

