## Supplemental Materials for "Real-time fMRI-informed self-regulation of the ventromedial prefrontal cortex using positive episodic future thinking enhances motivation"

**Supplementary Materials**

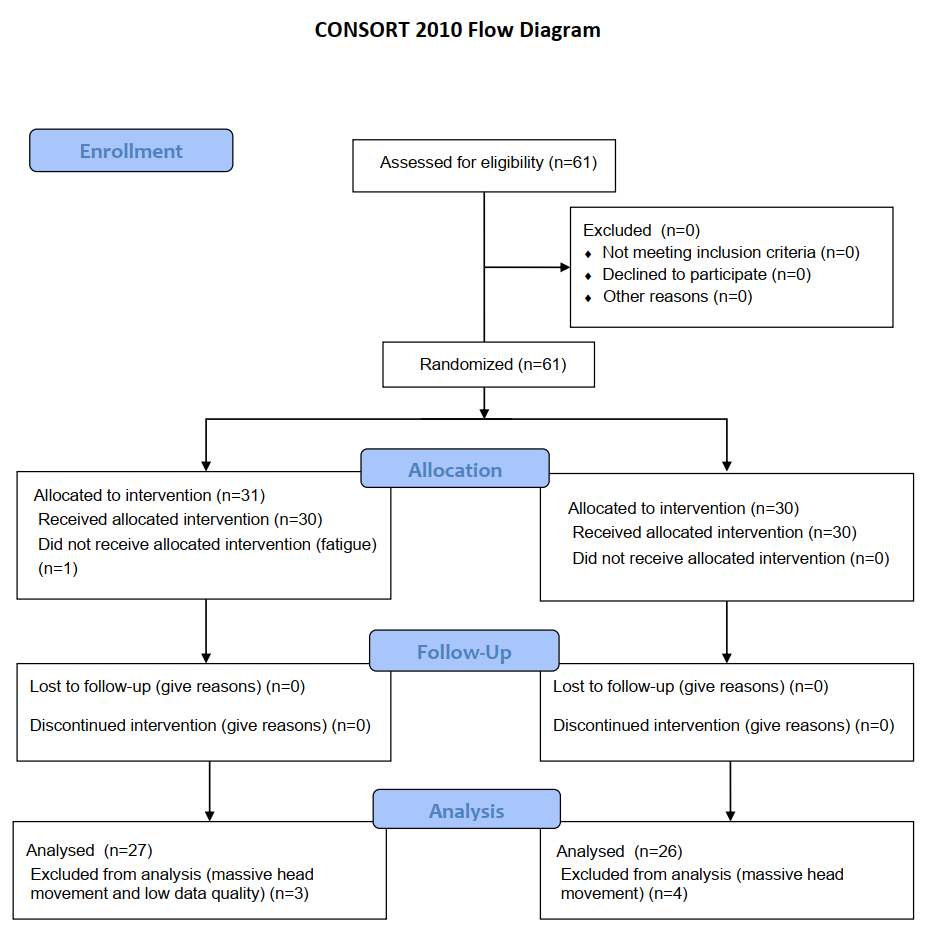

Figure S1. CONSORT flow diagram of the present study.

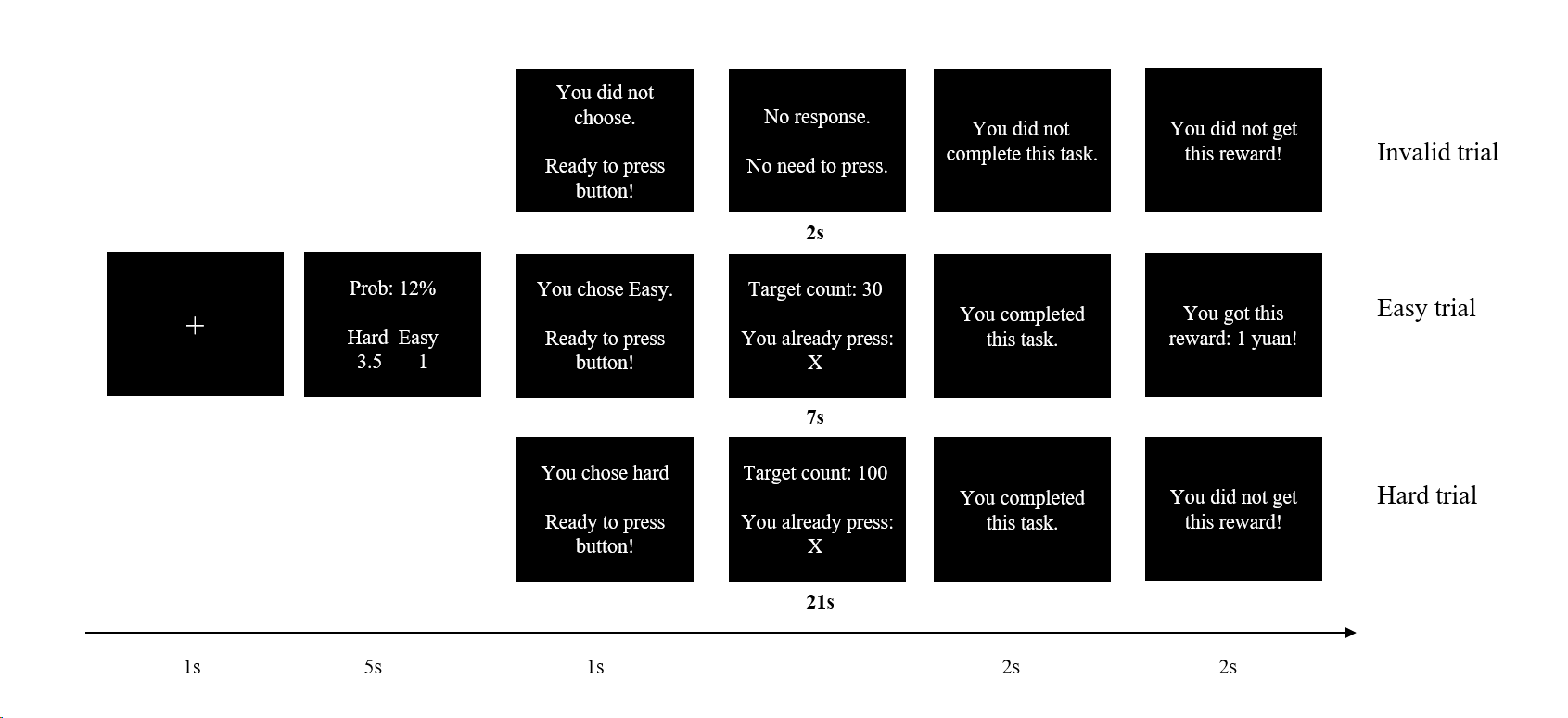

Figure S1. Experimental paradigm of the Effort-Expenditure for Rewards Task.

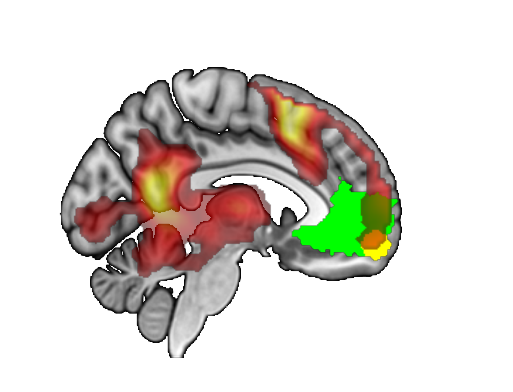

Figure S2. The overlap map between the ventromedial prefrontal cortex (vmPFC) and group maps in the baseline run. The green one is the anatomical mask of the vmPFC from the Yeo-17 atlas; The yellow one is the functional mask of the vmPFC which is created based on peak-coordinate in the vmPFC localizer task.

**Table S1**

Statistics of ages and questionnaire scores in the neurofeedback (NFT) and control (CTR) groups (mean ± SD).

| Questionnaires | NFT | CTR | t-value | p-value |
| --- | --- | --- | --- | --- |
| Number (Female) | 27 (12) | 26 (14) | / | / |
| Age | 21.37 (2.17) | 21.65 (2.12) | -0.481 | 0.632 |
| 1^st^-PAS | 26.48 (8.29) | 25.73 (5.40) | 0.389 | 0.699 |
| 1^st^-PNS | 13.96 (8.23) | 13.73 (5.60) | 0.12 | 0.905 |
| 1^st^-SAI | 38.44 (8.50) | 39.12 (9.63) | -0.269 | 0.789 |
| TAI | 41.44 (10.47) | 41.65 (8.61) | -0.079 | 0.937 |
| BDI | 8.63 (8.44) | 8.19 (7.58) | 0.193 | 0.848 |
| VVIQ | 60.89 (9.34) | 62.62 (9.90) | -0.653 | 0.632 |
| BIS | 15.26 (3.05) | 14.85 (2.88) | 0.507 | 0.614 |
| BAS | 22.07 (5.12) | 23.77 (3.44) | -1.419 | 0.163 |
| ZPTI | 16.62 (1.64) | 16.38 (1.24) | 0.611 | 0.544 |

*Note.* PANAS: Positive and Negative Affect Schedule; STAI: State-Trait Anxiety Inventory; BDI: Bermond-Vorst Alexithymia Questionnaire; VVIQ: Vividness of Visual Imagery Questionnaire; BIS/BAS: Behavioral Inhibition System and Behavioral Activation System Scale; ZTPI: Zimbardo Time Perspective Inventory.

**Table S2**

Statistics of questionnaire scores in the NFT and CTR groups before and after NF training (mean ± SD).

| Questionnaires | NFT | CTR | Group | Timepoint | Interaction effects |
| --- | --- | --- | --- | --- | --- |
| 1^st^-PAS | 26.48 (8.29) | 25.73 (5.40) | F(1,51)=0.339,  *p*=0.563 | F(1,51)=1.268,  *p*=0.265 | F(1,51)=0.219, *p*=0.642 |
| 2^nd^-PAS | 25.93 (9.73) | 24.38 (6.99) |  |  |  |
| 1^st^- PNS | 13.96 (8.23) | 13.73 (5.60) | F(1,51)=0.296,  *p*=0.588 | F(1,51)=6.129,  *p*=0.017 | F(1,51)=1.109, *p*=0.297 |
| 2^nd^-PNS | 10.81 (2.70) | 12.46 (4.97) |  |  |  |
| 1^st^-SAI | 38.44 (8.50) | 39.12 (9.63) | F(1,51)=0.178,  *p*=0.674 | F(1,51)=1.572,  *p*=0.216 | F(1,51)=0.117, *p*=0.734 |
| 2^nd^-SAI | 36.96 (9.26) | 38.27 (9.25) |  |  |  |

*Note.* PANAS: Positive and Negative Affect Schedule; STAI: State-Trait Anxiety Inventory;

**Table S3**

Brain regions showing brain activation in positive imagination relative to rest blocks in the vmPFC localizer run and tapping condition in the M1 localizer run across the 2 groups (MNI coordinates).

| Brain Region | BA | NO. Voxels | peak t-value | X | Y | Z |
| --- | --- | --- | --- | --- | --- | --- |
| **vmPFC localizer: positive imagination > rest** |  |  |  |  |  |  |
| L. Supplementary Motor Area | 6/8/32 | 1762 | 10.93 | -6 | 14 | 62 |
| Supplementary Motor Area |  |  | 10.68 | -6 | 14 | 50 |
| Middle Frontal Gyrus |  |  | 9.54 | -39 | 5 | 53 |
| Superior Frontal Gyrus |  |  | 5.98 | -9 | 56 | 35 |
| Inferior Frontal Gyrus |  |  | 5.18 | -45 | 20 | -10 |
| Ventromedial Prefrontal Cortex |  |  | 4.08 | -3 | 59 | -13 |
| Dorsal Anterior Cingulate Cortex |  |  | 3.67 | 9 | 20 | 38 |
| L. Posterior Cingulate Cortex | 30/31/39 | 3732 | 10.17 | -9 | -55 | 14 |
| Angular Gyrus |  |  | 6.16 | -45 | -67 | 26 |
| Thalamus |  |  | 6.10 | -3 | -13 | 5 |
| Parahippocampal Gyrus |  |  | 5.69 | -33 | -37 | -13 |
| Caudate |  |  | 5.59 | -18 | -1 | 20 |
| R. Middle Temporal Gyrus | 21 | 52 | 4.38 | 57 | -1 | -22 |
| R. Inferior Frontal Gyrus | 46 | 27 | 4.01 | 57 | 26 | -1 |
| Inferior Frontal Gyrus |  |  | 3.30 | 57 | 29 | 11 |
| R. Culmen |  | 20 | 3.68 | 18 | -40 | -28 |
| L. Middle Temporal Gyrus | 21 | 14 | 3.57 | -57 | -7 | -22 |
| L. Middle Temporal Gyrus | 22/39 | 50 | 3.32 | 39 | -55 | 20 |
| Middle Temporal Gyrus |  |  | 3.13 | 54 | -67 | 23 |
| R. Inferior Frontal Gyrus | 47 | 11 | 3.04 | 42 | 26 | -16 |
| **M1 localizer: tapping** |  |  |  |  |  |  |
| R. Lingual Gyrus | 6/18/40 | 17254 | 18.28 | 18 | -85 | -10 |
| Lingual Gyrus |  |  | 15.47 | -12 | -91 | -13 |
| Primary Motor Cortex |  |  | 13.09 | -39 | -22 | 50 |
| Thalamus |  |  | 12.29 | -12 | -22 | -1 |
| R. Postcentral Gyrus | 2/22/40 | 1473 | 10.28 | 60 | -31 | 44 |
| Superior Temporal Gyrus |  |  | 8.58 | 66 | -28 | 20 |
| Postcentral Gyrus |  |  | 8.29 | 57 | -19 | 20 |
| R. Middle Cingulate Cortex | 31 | 11 | 3.31 | 15 | -28 | 41 |

*Note.* All regions are reported with a p_FDR_ < 0.05 threshold at the whole-brain level. L indicates left; R indicates right.

**Table S4**

Brain regions showing stronger activity in regulation relative to rest blocks across training sessions in the NFT and CTR groups, respectively (MNI coordinates).

| Brain Region | BA | NO. Voxels | peak t-value | X | Y | Z |
| --- | --- | --- | --- | --- | --- | --- |
| **NFT group: regulation > rest** |  |  |  |  |  |  |
| L. Superior Frontal Gyrus | 6/10/32 | 532 | 7.77 | -9 | 14 | 56 |
| Superior Frontal Gyrus |  |  | 5.21 | -9 | 56 | 35 |
| Superior Frontal Gyrus |  |  | 4.63 | -15 | 47 | 41 |
| Ventromedial Prefrontal Cortex |  |  | 4.14 | -3 | 62 | -10 |
| L. Precuneus | 18/30/31 | 3400 | 7.55 | -24 | -52 | 8 |
| Precuneus |  |  | 7.06 | -18 | -46 | 5 |
| Posterior Cingulate Cortex |  |  | 6.79 | -9 | -55 | 14 |
| Thalamus |  |  | 4.53 | -15 | -13 | 2 |
| Caudate |  |  | 4.53 | -18 | -10 | 20 |
| Parahippocampal Gyrus |  |  | 3.99 | -21 | -25 | -16 |
| Hippocampus |  |  | 3.83 | 36 | -31 | -10 |
| L. Inferior Frontal Gyrus | 45/47 | 213 | 5.49 | -42 | 26 | -4 |
| Inferior Frontal Gyrus |  |  | 5.11 | -42 | 20 | -16 |
| L. Middle Frontal Gyrus | 6/8 | 128 | 5.47 | -39 | 5 | 56 |
| Thalamus |  | 54 | 3.49 | 15 | -22 | 20 |
| R. Extra-Nuclear |  | 14 | 3.46 | 21 | -16 | 8 |
| L. Middle Temporal Gyrus |  | 11 | 3.30 | -45 | -67 | 26 |
| R. Parahippocampal Gyrus | 35 | 10 | 3.10 | 24 | -19 | -16 |
| **CTR group: regulation > rest** |  |  |  |  |  |  |
| L. Supplementary Motor Cortex | 17/18/30 | 5033 | 7.63 | -6 | 14 | 59 |
| Superior Frontal Gyrus |  |  | 6.81 | -9 | 8 | 65 |
| Extra-Nuclear |  |  | 6.58 | -18 | -49 | 20 |
| Posterior Cingulate Cortex |  |  | 5.42 | -9 | -55 | 17 |
| Thalamus |  |  | 4.50 | -12 | -22 | 17 |
| Precunues |  |  | 4.32 | -3 | -61 | 20 |
| Caudate |  |  | 4.14 | 36 | -31 | -4 |
| Hippocampus |  |  | 3.82 | -36 | -28 | -13 |
| Parahippocampal Gyrus |  |  | 2.82 | 21 | -28 | -16 |
| L. Middle Frontal Gyrus | 6 | 96 | 4.41 | -36 | 2 | 50 |
| L. Superior Frontal Gyrus | 9 | 10 | 3.38 | -15 | 47 | 41 |
| Superior Frontal Gyrus |  |  | 3.34 | -15 | 53 | 35 |
| R. Extra-Nuclear |  | 25 | 3.37 | 21 | -7 | 26 |
| L. Superior Temporal Gyrus | 38 | 11 | 2.99 | -48 | 20 | -13 |

*Note.* All regions are reported with a p_FDR_ < 0.05 threshold at the whole-brain level. L indicates left; R indicates right. NFT: neurofeedback training; CTR: control.

**Table S5**

Regions that showed increased resting-state functional connectivity with the left vmPFC in Rest 2 relative to Rest 1 in the NFT and CTR groups, respectively (MNI coordinates).

| Brain Region | BA | NO. Voxels | peak F-value | X | Y | Z |
| --- | --- | --- | --- | --- | --- | --- |
| **NFT group: Rest 2 > Rest 1** |  |  |  |  |  |  |
| L. Dorsolateral Prefrontal Cortex | 10 | 34 | 20.61 | -45 | 48 | 0 |
| R. Posterior Insula | 13/41 | 51 | 19.17 | 48 | 0 | 3 |
| Posterior Insula |  |  | 17.51 | 48 | 6 | -9 |
| Posterior Insula |  |  | 13.44 | 42 | -6 | 6 |
| R. Declive |  | 21 | 19.12 | 12 | -78 | -21 |
| R. Uvula |  | 38 | 18.80 | 15 | -90 | -36 |
| L. Middle Cingulate Cortex | 24 | 14 | 18.34 | -9 | -21 | 42 |
| L. Posterior Insula | 13/22/42 | 121 | 17.38 | -42 | -9 | 9 |
| Posterior Insula |  |  | 17.07 | -42 | 6 | 3 |
| Posterior Insula |  |  | 17.04 | -45 | -3 | 3 |
| L. Angular Gyrus | 39 | 19 | 16.73 | -36 | -57 | 33 |
| L. Lingual Gyrus | 18 | 47 | 16.48 | -12 | -81 | -24 |
| **CTR group: Rest 2 > Rest 1** |  |  |  |  |  |  |
| R. Culmen |  | 54 | 21.91 | 9 | -54 | -30 |
| Culmen |  |  | 17.12 | 21 | -48 | -24 |
| L. Culmen |  | 20 | 18.74 | -15 | -45 | -27 |
| L. Superior Temporal Gyrus | 38 | 13 | 15.11 | -48 | 18 | -30 |
| Superior Temporal Gyrus |  |  | 14.61 | -45 | 12 | -42 |

*Note.* All regions are reported with a p_uncorrected_ < 0.001 threshold at the whole-brain level. L indicates left; R indicates right. NFT: neurofeedback training; CTR: control.
